# Cluster-randomized Trial of Homework, Organization, and Planning Skills Program Compared to Treatment as Usual/Waitlist for Youth Ages 11-14: Study Protocol for Conceptual Replication

**DOI:** 10.64898/2026.02.13.26346294

**Authors:** Jenelle Nissley-Tsiopinis, Phylicia F. Fleming, Wendy Chan, Joshua M. Langberg, Jaclyn Cacia, Tara Vigil, Brittany Chamberlin, Christina A. Di Bartolo, Katie L. Tremont, Emma H. Walz, Abbas F. Jawad, Jennifer A. Mautone, Thomas J. Power

## Abstract

**Background:** Organization, time management, and planning (OTMP) difficulties are associated with academic underachievement. OTMP skills training programs are effective in reducing OTMP deficits and improving academic performance. A randomized controlled trial of Homework, Organization, and Planning Skills (HOPS) for students ages 11-14 (1) found it to be effective with medium to large effects. In that study, HOPS was provided by counselors employed by the research team. This study is a replication examining HOPS under more authentic conditions when providers are employed by schools serving enrolled students. The primary aim is to evaluate HOPS offered by school providers in relation to treatment-as-usual/waitlist (TAU/WL). To respond to limited school resources post-COVID-19, HOPS is also provided by research team members, creating the opportunity to replicate the findings from the prior trial (1) and explore differential effectiveness when HOPS is implemented by school vs. research providers.

**Methods:** Students in about 30 schools serving students ages 11-14 will be enrolled. Schools are randomly assigned to HOPS vs. TAU/WL on a 2:1 ratio. Students assigned to HOPS schools are randomly assigned to a school vs. research provider on a 1:1 basis. Providers receive two hours of training and additional assistance on request. Child outcomes related to OTMP skills, homework, and academic performance are assessed at post-treatment, 6-month (from baseline) follow-up, and 12-month follow-up. HOPS sessions are video recorded for fidelity coding.

Potential effect modifiers include student ADHD, oppositional defiant, and internalizing symptoms, and family socioeconomic level. Analyses will use mixed effects modeling. The goal of the study is to enroll 135 participants, yielding a minimal detectable effect size of 0.50, within the expected range based on prior research.

**Discussion:** The study is unique in examining intervention implementation and effectiveness when intervention is provided under authentic practice conditions.

**Trial Registration:** This study was registered with clinicaltrials.gov (NCT04465708).

## Introduction

Among the many factors that influence academic performance, executive functioning (EF) deficits are key predictors (2). Organization, time management, and planning (OTMP) skills are aspects of EF that are particularly associated with students’ academic performance (3).

Organizational demands increase over the course of schooling and are highly salient during the middle school grades.

Given that OTMP skills are predictive of later academic achievement, schools need effective interventions to support students with deficits in these areas (3,4). Several intervention programs targeting OTMP skills have been developed and shown to be effective (1,5,6,7,8). For the middle school population (ages 11-14), the Homework, Organization, and Planning Skills (HOPS) program has been shown to be effective and feasible to implement. HOPS is a 16-session child skills training program that provides training to middle school students in school materials organization, homework management, task planning, and time management. HOPS was designed to be provided in 20-minute sessions on an individualized basis, although it can be implemented with a small group of students (9). In addition, HOPS includes parent consultation and often teacher consultation to promote generalization of newly acquired skills in real-world settings.

### Prior Research on HOPS Program

In an early study of HOPS, the program was implemented by school mental health professionals in participating schools to 47 students identified as having ADHD and academic problems (10). The study showed that school counselors demonstrated a high level of fidelity to intervention content. In comparison to a treatment-as-usual, waitlist (TAU/WL) control condition, caregivers in HOPS reported their children made significant improvements in organized actions (Cohen’s *d* = 0.88), task planning (*d* = 1.05), and homework completion behaviors (*d* = 0.85). Outcomes based on teacher ratings did not show improvement. Gains based on caregiver ratings generally were maintained at 3-month follow-up. In addition, HOPS participants demonstrated a superior grade point average (GPA) in comparison to TAU/WL.

HOPS was evaluated subsequently in a large-scale randomized controlled trial (RCT, *n*=280, 7 schools; (1)). The trial was conducted in authentic practice settings (schools). Intervention providers were school counselors who had recently graduated, but they were hired and supervised by the research team. The findings revealed that, compared to TAU/WL, students receiving HOPS demonstrated significant improvements with large effect sizes in caregiver ratings of homework performance (*d*=1.29), task planning (*d*=.79), materials management (*d* =.81), and organized actions (*d* = 1.14). Improvements based on teacher ratings were significant with medium effect sizes in materials management (*d* =.53) and organized actions (*d*=.55).

Findings showed evidence of sustained effects when assessed about 3 months after intervention. An examination of implementation factors revealed that counselor-reported working alliance was a significant predictor of favorable effects of HOPS on outcomes (11).

### Gaps in Research on HOPS Program

Although the HOPS program was designed primarily for students with ADHD, it has the potential to be effective for students with OTMP deficits who do not have this disorder (9).

Indeed, HOPS may be more useful to school professionals if it can be provided to the broader population of students with OTMP problems, not just those with ADHD (8). Deficits in OTMP skills are highly common among students with ADHD (12), and they also occur among students with autism spectrum disorder (13), learning disabilities (14), and adverse psychosocial childhood experiences (15). As such, this study will include students in general education classrooms demonstrating OTMP deficits that may be impacting their academic functioning, regardless of diagnostic status.

HOPS is unique among psychosocial interventions in that it was developed in partnership with schools with a focus on provider usability and feasibility of implementation. However, the manner in which HOPS was applied in the large-scale RCT differs in some important ways from how it likely is applied in routine practice conditions. When schools make the choice to implement HOPS, they need to make use of existing resources. For example, school administrators need to identify one or more school staff to provide the intervention to students.

Research is needed to inform school administrators about whether HOPS works in routine practice when the program is offered by school staff. In the original design of the current study, the plan was for HOPS to be provided solely by school professionals working in the school.

Because the initial years of this study were implemented during the COVID-19 pandemic, which placed limits on the capacity of schools to participate in this trial, the study design was modified in response to input from school professionals and funding sponsors so that HOPS will be provided by both school professionals (school team providers) as well as educational professionals contracted by the research team who do not work in the schools (research team providers). This shift in design affords the opportunity to test the effectiveness of HOPS when delivered under conditions similar to the original trial (i.e., implementation by members of the research team, (1) as well as conditions more similar to actual practice (i.e., implementation by school providers).

### Importance of Intervention Implementation and Relational Processes

Intervention effectiveness depends to a substantial extent on how well the intervention is implemented (provider fidelity) and the extent to which participants are engaged or involved in the intervention (16,17). Provider fidelity refers to adherence to the steps of the intervention curriculum as well as level of competence in providing intervention. Participant engagement includes attendance, active participation in sessions, and adherence to recommended intervention practices (16). Another important implementation factor is the working alliance between the provider and student, or the ability of the provider to work with the student in a responsive, collaborative manner (18). Monitoring implementation processes (provider fidelity, student and parent engagement) and assessing working alliance are especially important when interventions are provided under routine practice conditions, because there is less control over provider fidelity and often less emphasis on promoting intervention in a replicable, standardized manner. In this study, our team will carefully examine provider fidelity, student and parent engagement, and working alliance between the intervention provider and student participants and evaluate the extent to which these factors are associated with improvements in student outcomes.

### Conceptual Framework of HOPS

The HOPS program is based primarily on principles of behavior therapy and incorporates elements of cognitive behavioral therapy. Behavioral strategies include verbal instruction, prompting skill use, shaping, and contingency management (with an emphasis on positive reinforcement for effort and performance of targeted OTMP behaviors). HOPS includes parent consultation, and often teacher consultation, to foster generalization of skills across settings and maintenance of strategy use after the intervention has ended. In addition, cognitive-behavioral strategies are used to identify and modify negative, defeatist attitudes that prevent the child from making progress (9).

Several factors, including provider intervention fidelity, as well as student and caregiver engagement in HOPS likely contribute to changes in students’ OTMP skills and homework performance (proximal outcomes). In turn, based on previous research (19,20) changes in OTMP skills likely lead to improved academic (distal) outcomes. Based on research from the previous evaluation of HOPS (1), we expect that severity of student organization problems, ADHD symptoms, and oppositional defiant behaviors at baseline will moderate the effect of intervention on OTMP skills and homework performance. In addition, based on prior research on child organizational skills training at the elementary level, we expected severity of child internalizing symptoms (anxiety and depression) baseline will moderate the effect of intervention (20). We will also examine whether child medication status (i.e., taking or not taking medication to treat ADHD) and family socioeconomic status modify the effect of intervention. (See Figure 1 for the Spirit Checklist and Figure 2 for the HOPS theory of change.)

**Figure 1:**
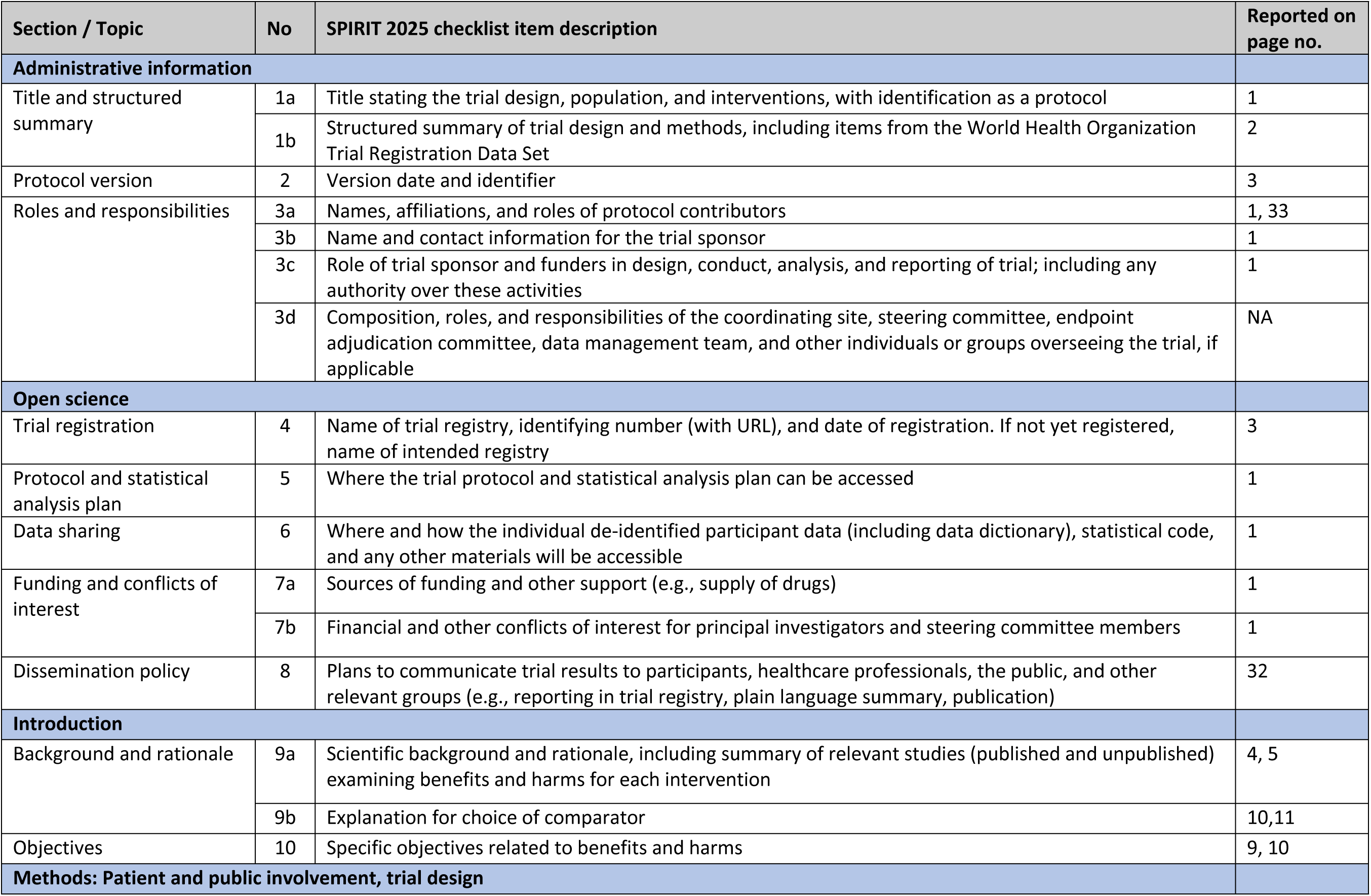
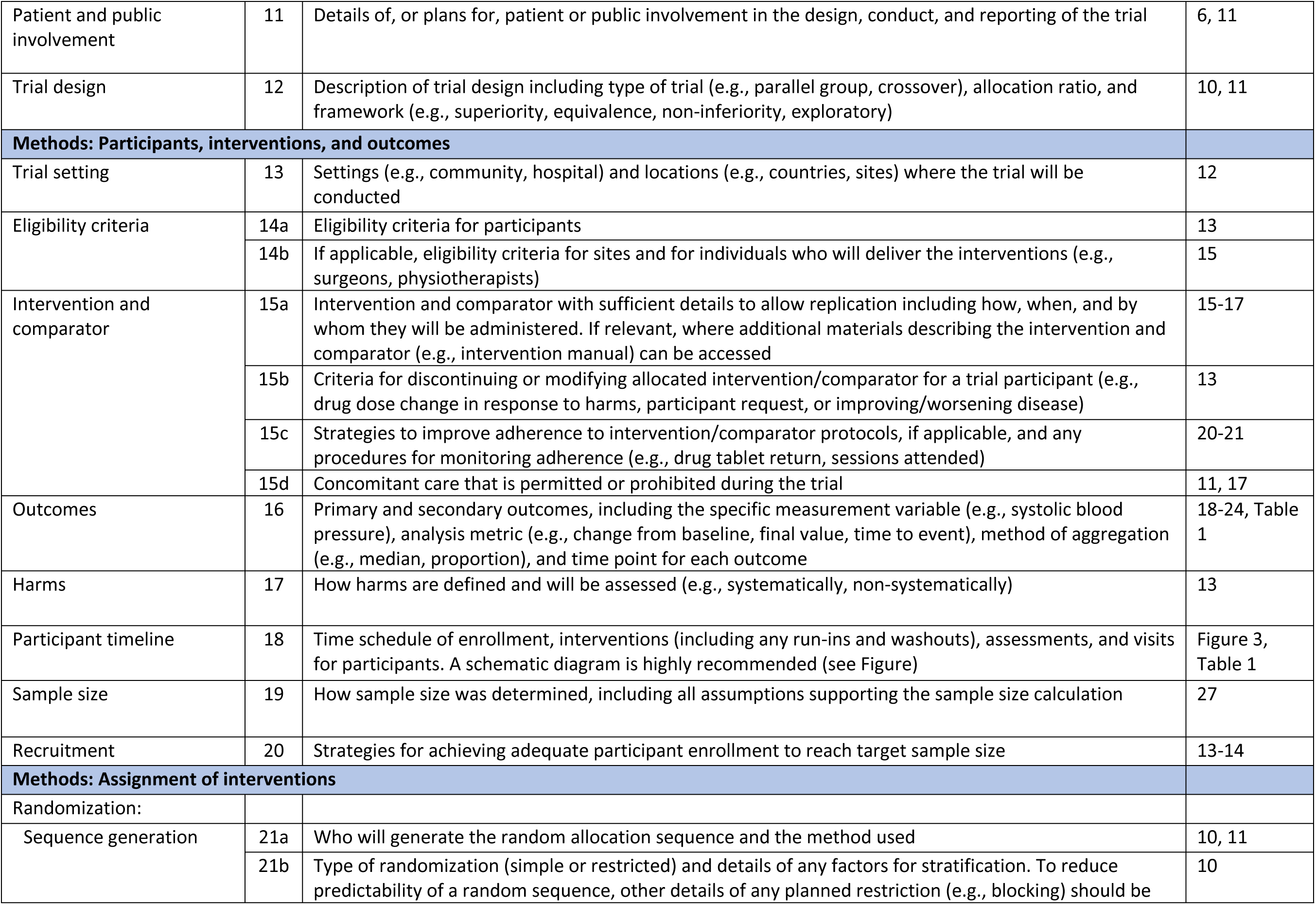
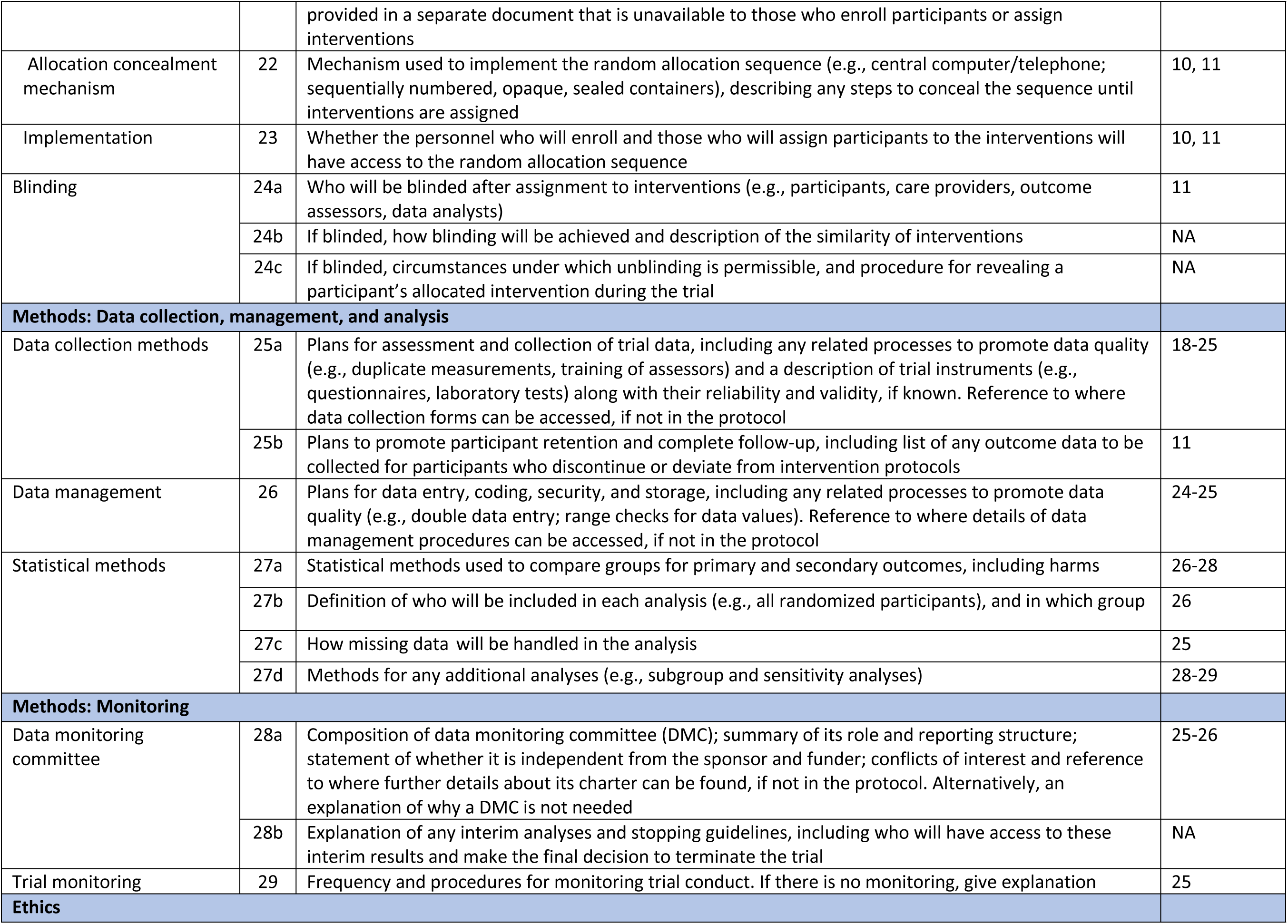
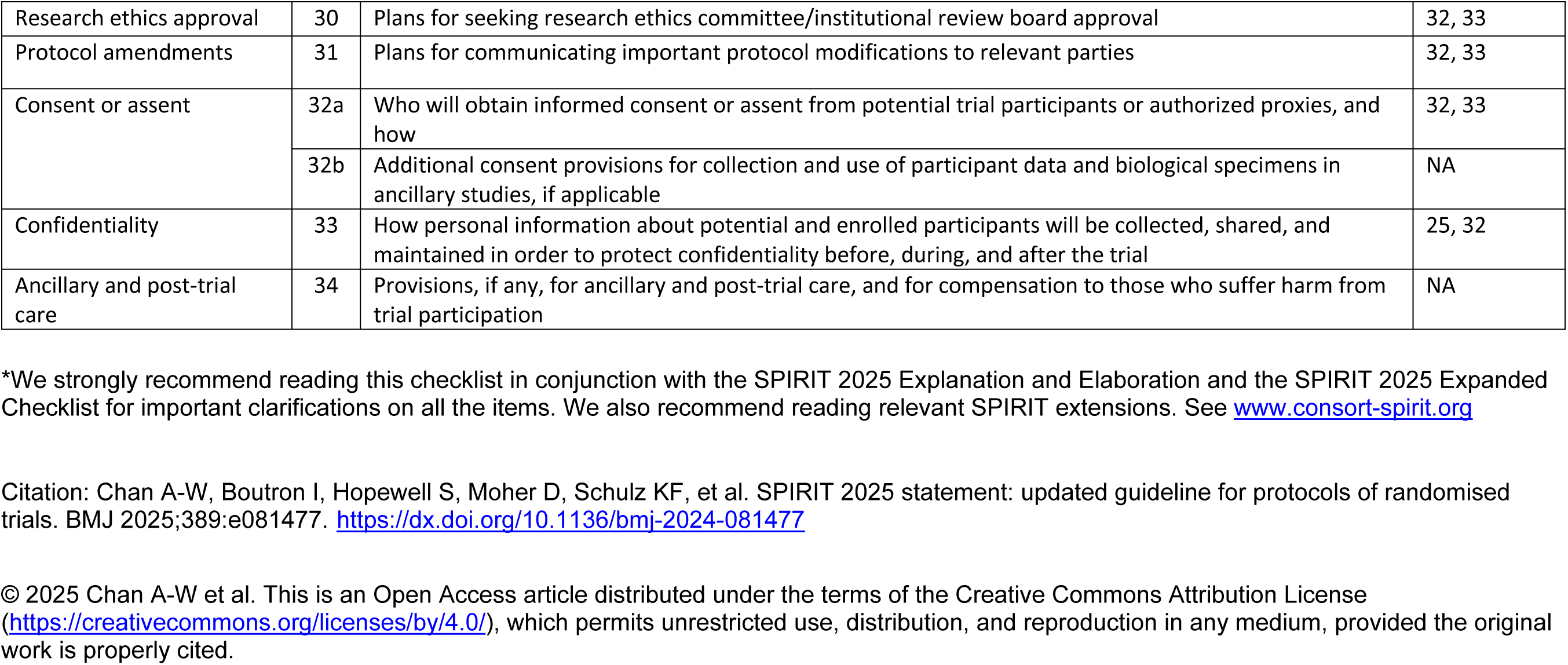
SPIRIT 2025 checklist of items to address in a randomized trial protocol for the HOPS study**\***

**Figure 2.**
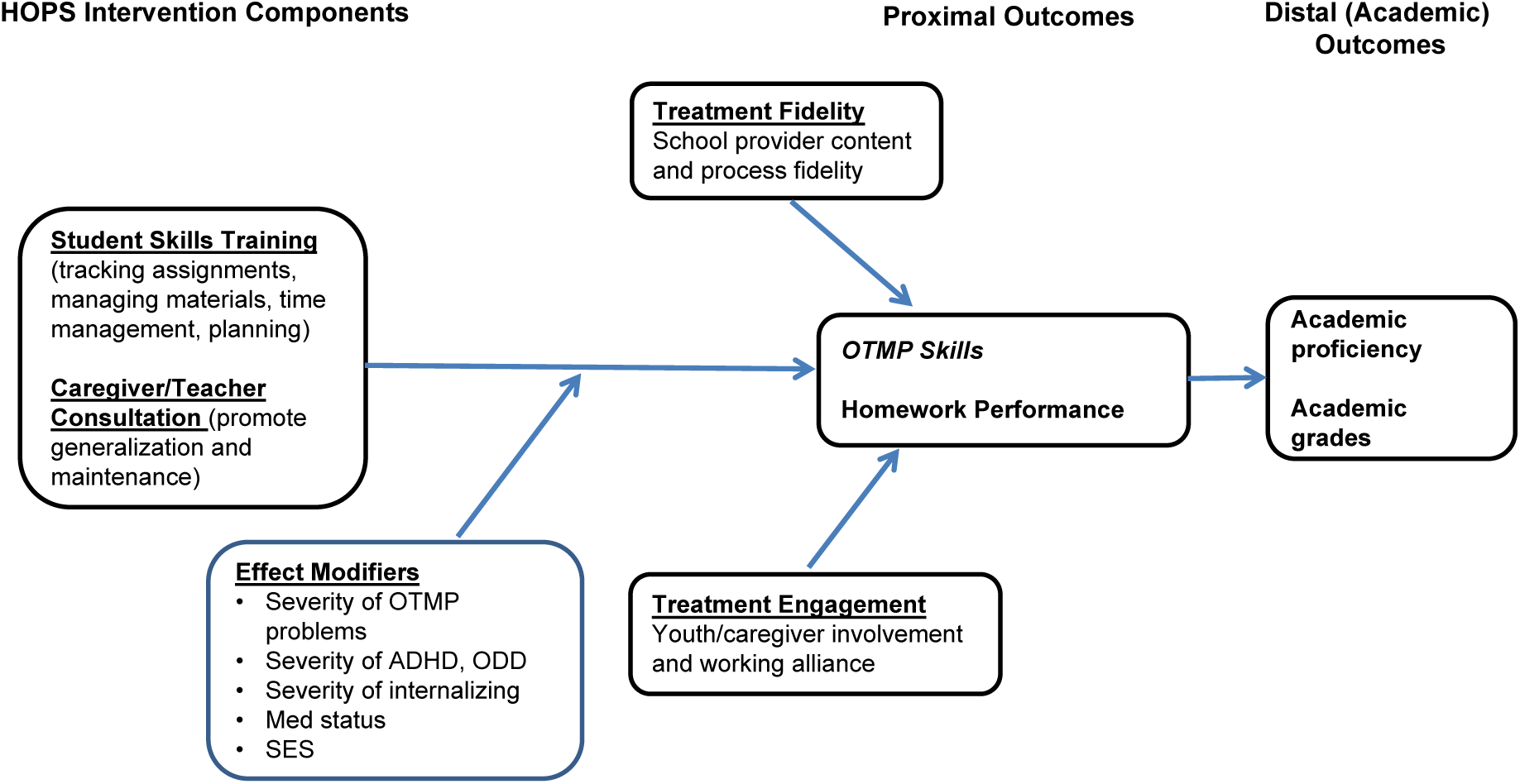
HOPS study theory of change. The figure depicts HOPS intervention components and factors that may impact the effect of HOPS on proximal and distal outcomes. OTMP = organization, time management, and planning skills; ODD = oppositional/defiant behavior; SES = socioeconomic status

### Study Purpose and Aims

The purpose of this study is to conduct a replication (i.e., replication with design elements that differ from the original trial) of a randomized controlled trial that showed the HOPS intervention provided by counselors employed by the research team improved organizational, time management, and planning (OTMP) skills for middle school students diagnosed with ADHD in relation to students on a waitlist receiving treatment-as-usual (TAU/WL; (1)). The trial will be conducted by an independent research team; the developer, Dr, Langberg, and his team will provide training, technical assistance and follow-up consultation on request to HOPS providers but will not be involved in any intervention evaluation activities.

Similarly, the research team evaluating the intervention will not be involved in training or supervising HOPS intervention implementation. The study will test whether HOPS compared to TAU/WL yields similar benefits when implemented by providers on the research team (HOPS-Research Team; HOPS-RT) and school-based professionals (HOPS-School Team; HOPS-ST) for the full range of students who have OTMP skills deficits, not only those with a formal diagnosis of ADHD. The study will examine six aims:

**Aim 1:** Evaluate the difference between HOPS-ST and TAU-WL on improvement in proximal outcomes (organization skills, homework performance) and distal outcomes (academic performance) at post-treatment (4-months after baseline) and explore the intervention’s sustainability (6 and 12-months after baseline). This aim will enable our team to determine the effectiveness of HOPS when provided by school professionals. We hypothesize that students in the HOPS-ST group will demonstrate significantly fewer OTMP skills deficits and homework problems and will show significantly better academic performance at post-treatment compared to students in TAU-WL. In addition, we expect the benefits of HOPS-ST to be maintained at follow-up.

**Aim 2:** Examine the effectiveness of HOPS-RT in relation to TAU-WL. This aim will enable our team to replicate the findings of the Langberg et al. (1) trial. We hypothesize that students in the HOPS-RT group will demonstrate significantly fewer OTMP skills deficits and homework problems and would show significantly better academic performance at post-treatment compared to students in TAU-WL. In addition, we expect the benefits of HOPS-ST to be maintained at follow-up.

**Aim 3:** Explore differences in effect for HOPS-ST and HOPS-RT at post-treatment and follow-up.

**Aim 4:** Explore mechanisms of action, specifically whether improvements in OTMP skills mediate the effect of HOPS on improvements in student homework and academic performance. **Aim 5:** Explore whether student grade level, severity of organization problems, severity of ADHD, oppositional defiant behavior, and internalizing symptoms, and student medication status at baseline moderate the effect of HOPS on outcomes.

**Aim 6:** Explore the extent to which treatment fidelity, student engagement, parent engagement, teacher engagement, and provider-student working alliance predicts outcomes for students in HOPS.

**Aim 7:** Estimate the cost of providing HOPS under authentic practice conditions in schools.

## Materials and Method

### Trial Design

#### Random Assignment to Study Conditions and Providers

The study is a two-group, parallel randomized controlled trial that is designed to include about 30 schools. Random assignment will be conducted at the level of schools and will occur after the school grants approval and the school provider offers consent. The study has two arms: HOPS and TAU-WL. Schools will be stratified by subsidized lunch status (low, medium, high) and randomly assigned to HOPS or TAU-WL at a 2:1 ratio. The sequence of assignments to treatment conditions will be determined using a computer-generated list of random identification numbers, conducted by the study statistician to ensure an unbiased and unpredictable assignment of participants to the study arms. The list of random numbers is generated using the statistical software program R (21). The study team will inform the principal and school provider after random assignment of the school to study condition has been made. The study is designed to compare HOPS to what students typically receive to address their OTMP skills deficits. A WL design was chosen to be responsive to feedback from school officials who expressed concerns about participating in the study if identified students were not able to obtain HOPS. Students in TAU/WL schools will receive HOPS later in the school year of enrollment or in the school year following the year of enrollment. Students enrolled in the highest grade level in the school (e.g., 8^th^ grade) will be enrolled in the first few months of the school year, so that there is enough time for them to receive HOPS before the end of the school year if their school is assigned to the TAU/WL condition.

#### Random Assignment of Students in HOPS Schools to School or Research Provider

Students randomly assigned to the HOPS condition will be randomized once again to receive HOPS from either a school provider or research provider (on a 1:1 basis). The sequence of assignment of students in the HOPS condition to the school or research provider will be determined by a separate computer-generated list of random identification numbers overseen by the team statistician. Caregivers and teachers will be informed about study assignment after caregiver consent and child assent is provided.

#### Concealment

Given that the study is a pragmatic trial being conducted under real-world conditions, no attempt will be made to conceal treatment assignment from caregivers, students, teachers, and intervention providers. In addition, study team members will be fully aware of treatment assignment.

#### Collection of Outcome Data

Outcome data for students in HOPS and TAU-WL will be collected at baseline and post-treatment (about 4 months after baseline). Follow-up data will be collected 6 months after baseline for students assigned to the HOPS condition. In addition, follow-up data will be collected 12 months after baseline for students in HOPS who remain in their school of enrollment the following school year. We will not collect 6-month and 12-month follow-up data for students in TAU-WL because of ethical concerns about further delaying the delivery of HOPS to those in the control group. However, we will collect post-treatment outcome data after students in the TAU/WL condition receive the HOPS program. Treatment fidelity, student engagement, and parent engagement will be assessed for participants in the HOPS condition during the intervention period. Provider-student working alliance will be assessed after the intervention period. Severity of organization problems, ADHD symptoms, disruptive behavior symptoms, and anxiety symptoms, as well as school organizational factors that may contribute to intervention implementation, will be assessed at baseline. (See Figure 3 for the study plan and flow diagram and see Table 1 for a list of measures and the timelines for administering measures).

**Figure 3.**
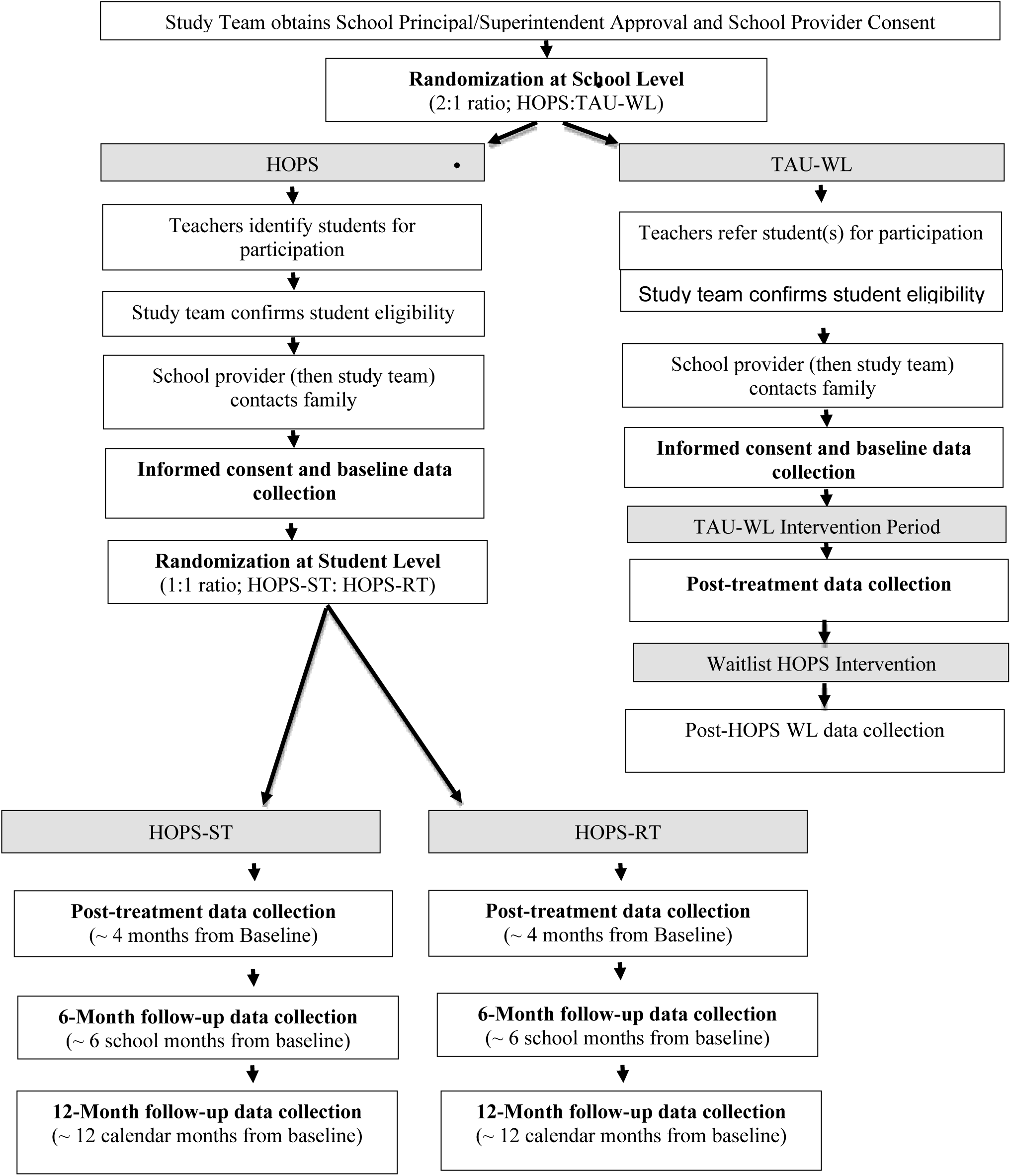
Study plan and flow diagram. The figure depicts random assignment to HOPS and TAU/WL, random assignment to school vs. research provider for participants in HOPS, and data collection at four time periods. HOPS-ST refers to HOPS implemented by a school team member, and HOPS-RT is HOPS implemented by the research team.

**TABLE 1:**
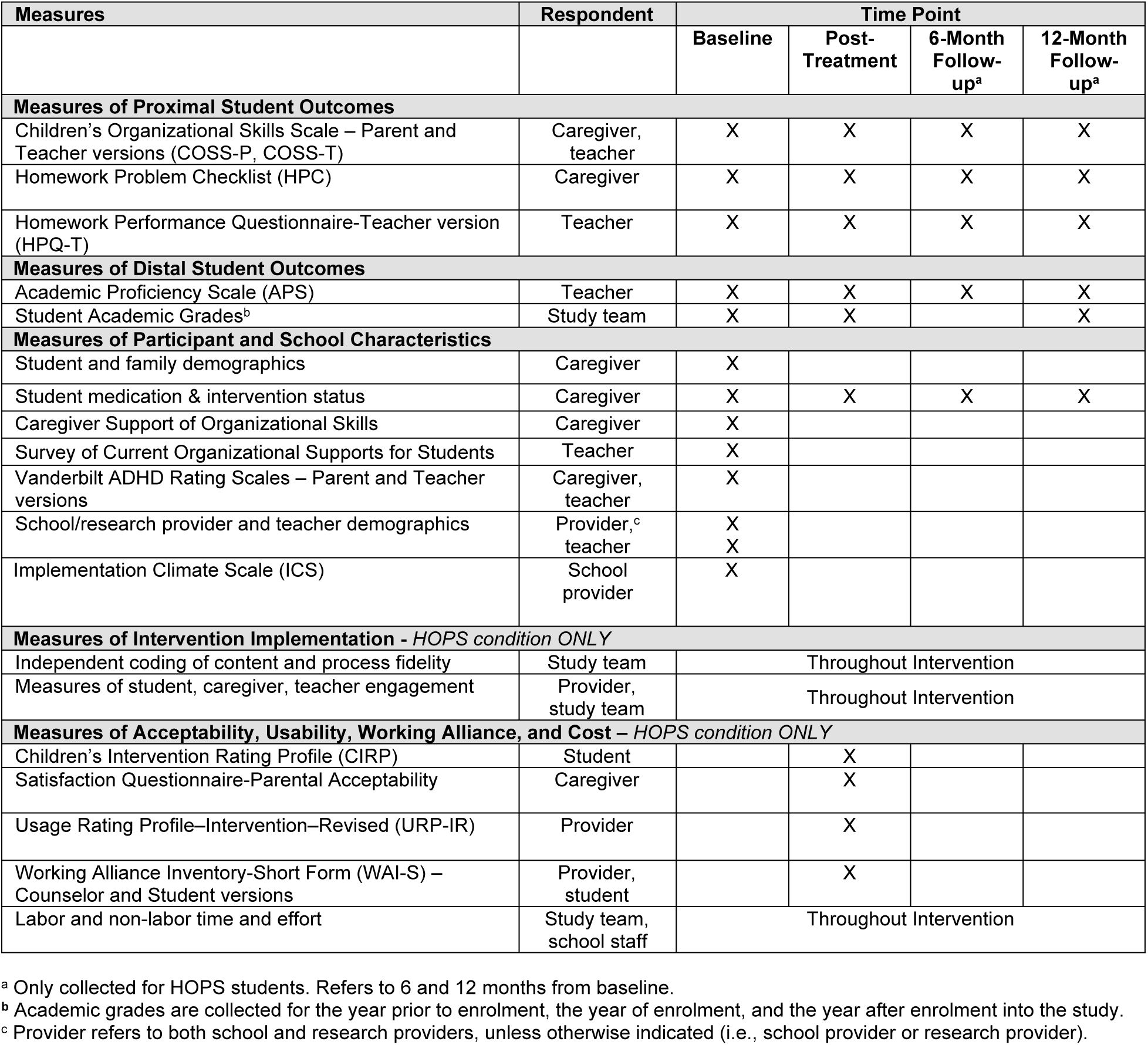
TIMELINE OF STUDY EVALUATION AND MEASUREMENT.

### Research Settings

The study will be conducted in Philadelphia, PA greater metropolitan region. Our team will purposely recruit schools that are highly diverse in subsidized lunch rates of the schools and racial and ethnic composition of the students to maximize generalizability. Our team will attempt to recruit schools with varying levels of urbanicity (i.e., rural, suburban, and urban). We expect the racial and ethnic minority composition across schools to range from 15% to 99% and subsidized lunch rates to range from 15% to 100%. For each school, we plan on recruiting one or two school providers to implement the HOPS intervention.

### Student Participants

Students will be eligible for this study if: (a) they attend a school participating in the study, (b) they are enrolled in grades 6 through 8 and placed in general education for a majority of the day, and (c) they are identified as having OTMP deficits that are impairing academic performance, as identified by the four interference items on the Children’s Organizational Skills Scale-Teacher version (COSS-T; (22)). Students classified with emotional/behavioral disorders and/or learning disabilities will be included if it is determined that the students’ disabilities are being addressed adequately in their current educational program and OTMP skill deficits are a primary concern. Students with a one-to-one aide will be excluded because the presence of an aide substantially alters how an organizational intervention is implemented. In addition, students from families in which both caregivers do *not* speak English will be excluded because the program has not yet been developed for non-English speakers. Parents or legal caregivers must give consent, and students must provide assent for student and family participation, which can be withdrawn at any time during study participation. The potential for harm to participants is minimal, but they are informed that it is possible they may feel uncomfortable in completing measures. Participants are not mandated to complete items on measures used in the study.

### Recruitment

Two primary methods will be used to recruit schools. First, emails will be sent to the principals of schools serving students in grades 6 to 8 located in the large metropolitan area in which the study will be conducted. The selection of schools will be inclusive of districts across the region to achieve balance across subsidized lunch rate levels of the schools (low [0%-30%], middle [31%-60%], and high [61%-100%]). In addition, there will be an intentional effort to recruit schools in urban, suburban, and rural areas. Second, emails will be sent to principals of schools from districts in which we have previously conducted research or those referred to us by school administrators or other researchers. Approval will be obtained from the school principal, school superintendent, and school board (as necessary) prior to conducting research in the school. In addition, principals will be asked to identify one or more school providers to implement the HOPS intervention. School provider(s) must provide consent to participate in the study for the school to be enrolled in the study. Once all approvals and consents are obtained from school professionals, the school will be randomly assigned to a study condition (HOPS vs. TAU/WL).

Students will be referred to the study by their classroom (general education) teachers.

School principals will identify the specific strategies to be used in convening teachers, informing them about the study, and obtaining referrals. Teachers will be informed that referral of a student to the study indicates their willingness to complete forms for outcome evaluation. The pool of teachers selected for inclusion in the referral process will be asked to nominate students from their general education classroom: (a) who are struggling the most with OTMP skills, (b) whose OTMP skill deficits interfere with their academic performance, and (c) who have at least one caregiver who speaks English. Each teacher who agrees to participate will complete a nomination form for each referred student for the purpose of determining student eligibility.

After a student is referred and determined to be eligible, a professional from the school, typically the school provider, will attempt to contact the parent or legal guardian to explain the study, indicate their child has been referred for HOPS through the study, and determine if the family is willing to be contacted by the study team. The school professional will often send a letter to the parent/guardian that includes a link to an electronic survey to offer permission for the study team to contact the family and provide contact information. The school provider may also contact (phone or email) parent(s)/guardian(s) of identified students to introduce the study. When the school gets permission from a family to share contact information with the study team, they will pass that information to the study team. With caregiver approval, a member of the study team will contact the family to determine interest, obtain caregiver consent, and obtain student assent.

### Intervention Conditions

The study has two intervention conditions: HOPS intervention implemented by a research or school provider and TAU/WL.

### HOPS Program

HOPS includes 16 sessions with the student, each about 20 minutes long. HOPS has three components: (1) materials organization, (2) homework management, and (3) time management. In addition, the program includes two caregiver consultations and, when feasible, one teacher consultation to inform them how they can support implementation of HOPS strategies at home and school. In this study, HOPS will be implemented by a school provider or research provider. The school provider can be a teacher (general or special education) or member of the student support staff (e.g., school counselor, school psychologist). The research provider will be a member of the research team with experience in special education, school psychology, counseling, or education research.

#### Materials Organization Component

Developing an efficient and structured system for managing school materials is a major goal of HOPS. During the first week of the program, students work with their provider to establish a system of backpack, binder, and computer files organization. An organization checklist, consisting of operationalized criteria for backpack, binder, and computer files organization is utilized. Baseline organization is recorded on the first session, and an organization system based on the checklist is established during the second session. The organization checklist is completed at every following intervention session during the program. Intervention providers work with students to develop strategies for maintaining their materials organization system.

#### Homework Recording and Management Component

The goal of this component is for students to establish a structured system for keeping track of and managing homework assignments. Students use a planner or calendar where they record all of their homework assignments and upcoming tests. Even if this information is available on different teacher websites, students transfer that information to their planner so they can see it all in one place and learn to plan ahead. Accuracy of recording is checked and documented by the provider at each HOPS session. Students also maintain a homework folder in their school binder. All assignments to be completed or papers to be signed are placed on the left side of the folder, and all completed assignments to be turned in are placed on the right side. As part of the organization checks, intervention providers evaluate use of the homework folder to ensure that only current assignments are in the folder and all other work is filed in the appropriate section of the binder.

#### Time Management Component

The goal of this component is for students to reduce procrastination by planning more effectively for the completion of long-term projects and tests. A time management skills checklist is utilized to guide implementation of time management strategies. The time management skills checklist includes operationalized criteria separated into two categories: (1) planning for upcoming tests and quizzes and (2) planning for projects. A tiered approach is used to ensure students master the most basic skills (e.g., recording a test in the planner) before attempting more complex skills (e.g., breaking test studying into smaller pieces).

#### TAU/WL

Students in the TAU/WL condition will receive the supports for OTMP skills typically offered to students in their schools. Teachers will be asked to complete a questionnaire at baseline to determine the OTMP supports being received by each student. Students in this condition will receive the HOPS program from a school or research provider after post-intervention data have been collected, which could be in the spring of the year of enrollment or the following school year.

### Training and Technical Support for HOPS Providers

Procedures for offering training and technical support for intervention providers were adapted from those used in prior studies of HOPS (1,10) and reflect the strategies used in actual practice by Langberg and colleagues when school professionals request training to implement HOPS. School and research providers will receive a copy of the HOPS manual (9) and 1.5-2 hours of training before providing HOPS. The training will include a description of: (a) the conceptual model and core principles of HOPS, (b) principles for promoting behavior change, (c) major components of the program, (d) procedures for providing each student session, (e) procedures for providing parent consultation (and teacher consultation when feasible), and (f) guidelines for selecting reinforcers for students. The training is provided virtually by Dr.

Langberg or a trainee working under his direction.

Intervention providers will be able to login to a website (hopsintervention.com) that will give them access to videos of Dr. Langberg providing HOPS training and responding to common treatment challenges, access to PowerPoint slides for Dr. Langberg’s training, and responses to Frequently Asked Questions (FAQs). Through the website, intervention providers will be able to ask brief logistical/technical questions about HOPS, which will be answered regularly by the training team (Dr. Langberg and trainees). In addition, intervention providers will be given the opportunity to receive consultations on demand via telephone, virtual communication, or email with this trainee. The training team will log each consultation call to record the name of the provider, topics discussed, and duration of communication.

### Outcome Measures – Proximal and Distal

The *Children’s Organizational Skills Scale – Parent and Teacher versions (COSS-P, COSS-T)* will be used to assess changes in OTMP functioning at home and school. COSS Total scores (26 items for COSS-P, 28 for COSS-T) have strong construct and discriminant validity (23) and treatment sensitivity (1). Each item is rated on a 4-point scale. The total score will be used in analyses, with higher scores being less favorable.

The *Homework Problem Checklist (HPC)* is a 20-item caregiver-report measure that indicates the frequency of child homework problems. The HPC has been shown to have strong construct validity (24) and treatment sensitivity (1). Each item is rated on a 4-point scale. The total score will be used in analyses with higher scores indicating more problems.

The *Homework Performance Questionnaire – Teacher version (HPQ-T)* assesses students’ homework behavior during the past 4 weeks. Each item is rated on a 7-point scale indicating how often each behavior occurs, with higher scores indicating better performance. The 17-item General factor that assesses overall homework competence has strong construct validity (25) and will be used in analyses.

The *Academic Proficiency Scale (APS)* is a teacher-report measure assessing proficiency in six academic subjects. Each item is rated on a 5-point scale (1=well below standard expected at this time of year; 3=at standard; 5=well above standard). This measure has high internal consistency (8) and strong treatment sensitivity (5). The mean item score will be used in analyses, with higher scores indicating better performance.

Student *academic report card grades* will be used to examine the impact of HOPS vs.

TAU/WL on academic performance. Student grades will be recorded for the last marking period of the year prior to enrollment (baseline), the year of enrollment (post-treatment), and the year following enrollment (follow-up). Grades will be harmonized across participating school districts to a uniform grading scale (ranging from a low of 1 to a high of 4). A composite academic grade score will be computed for each student based on their grades in math and reading/language arts (possible range = 1 to 4). Higher scores indicate better outcomes.

### Participant, Family, and School Characteristics

Caregivers will report information about student age, gender, race and ethnicity, caregiver education, family composition, and academic support services received by students outside of school. School staff will report on students’ special education status, amount of special education services received, Section 504 services, and other services being provided to address student organizational problems. Publicly-available, online data about the free and reduced-price lunch percentage of each school will be accessed. A geo-mapping tool (Area Deprivation Index, ADI,(26)) will be used to determine each family’s neighborhood level of disadvantage; higher scores on this measure indicate greater disadvantage.

Medication and educational intervention status will be assessed at baseline and each assessment point by asking caregivers to indicate whether the child is currently on psychotropic medication, name of medication, and date of initiation. We will monitor change in medication status over the course of the intervention and follow-up periods. Caregivers will also be asked to indicate whether their child is receiving intervention (e.g., therapy, skills training, coaching) related to organizational skills/executive functioning.

Caregivers will report on their support of their child’s organizational skills related to materials management, homework management, and planning (i.e., how much time and support they provide to the child to maintain these skills).

The *Vanderbilt Rating Scales* will be used to assess severity of ADHD, oppositional defiant behavior, and internalizing symptoms. The caregiver-and teacher-report versions of the Vanderbilt scales will be used (27,28). Informants rate how frequently each symptom occurs on a 4-point Likert scale (0=never, 3=very often). Both scales consist of a 9-item Inattention factor, a 9-item Hyperactivity-Impulsivity factor, an 8-item Opposition-Defiant factor, and a 7-item Internalizing factor. The internal consistency of each of these factors with middle school students has been shown to be high (>.90)(1).

HOPS providers, including school and research providers, will report on their professional roles, years of experience, gender, race and ethnicity, educational level, and years of experience. School providers will report aspects of the organizational (school) context that are relevant for the implementation of evidence-based practices in schools. The *Implementation Climate Scale* has 15 items; each item is rated on a five-point Likert scale (0=not at all; 4=very great extent). Higher scores indicate a more favorable climate. Research has supported the factor integrity and internal consistency of this scale (29).

### Measures of Intervention Implementation

#### Assessment of Content and Process Fidelity

The study team will video record intervention implementation for all 16 student sessions using a HIPPA compliant Outlook Teams meeting link; intervention providers will be asked to audio record each of the student sessions as a backup. Videotaped sessions determined to be key to intervention implementation because a new checklist is introduced (Sessions 2, 4, 8, 12), as well as the last self-management problem solving content session (Session 14) will be coded for each participant. In addition, one randomly selected session will be coded for each participant. In total, implementation fidelity will be coded for 6 of 16 student sessions (37.5%) to capture variations in implementation fidelity. Further, the study team will attempt to video record the caregiver and teacher sessions whenever possible, with intervention providers asked to audio record them as backup, and members of the research team will code a randomly selected subset of these sessions for fidelity. To evaluate the reliability of fidelity coding, a second, gold standard coder will double-code 25% of randomly selected, coded sessions.

The research team will evaluate content fidelity (treatment adherence) and process fidelity (intervention provider competence). Following the procedures used by Langberg and colleagues in their RCT (1) but modified slightly to account for degree of implementation, content fidelity items will specify critical steps of the intervention and each item will be rated as a 0, 1, 2, or 3 (0 = not implemented; 1-2 = partially implemented; 3 = fully implemented). Scores on each item will be collapsed to indicate whether an item is implemented (2 or 3) or not implemented (0 or 1). In addition, scores will be recorded on a four-point scale to indicate the extent to which content was implemented. Process fidelity (competence) will be rated on a scale from 1 to 4 (1 = rarely; 4 = all or almost all of the time; adapted from Dumas et al. (30)). The process fidelity items will include those we have identified as important to organizational skills training interventions in our currently funded study of organizational skills in elementary school (e.g., using positive reinforcement for skill implementation, engaging student in practice and implementation planning, using a growth mindset rather than correction to provide feedback for skill use, individualizing content to student’s situation, and presenting key content clearly).

Process fidelity will be captured via two forms: Process Fidelity Form for HOPS Student Sessions and Process Fidelity Form for HOPS Caregiver/Teacher Sessions.

#### Assessment of Intervention Engagement

Intervention providers will record student attendance at HOPS sessions and the duration of each session. The intervention provider will rate how involved the student was during and between sessions on a 6-point Likert scale (0 = not at all true to 5 = very true) (adapted from Chu & Kendall (31)). The average involvement score across sessions will be used as the index of student engagement. In addition, intervention providers will record caregiver attendance at caregiver consultation sessions. Using a measure developed by Langberg and colleagues (1), and adapted by the current study team,the HOPS Study: Caregiver Engagement Rating Scale, the intervention provider will rate how engaged the caregiver was during each meeting using a scale similar to that used for students. Further, at the second caregiver meeting intervention providers will rate the extent to which the caregiver seems invested in implementing the monitoring and reward plan they had created using a 5-point Likert scale (1 = not at all to 5 = very much). A similar process is used to assess teacher engagement using the HOPS Study: Teacher Engagement Rating Scale. The reliability of these ratings will be determined by having study staff code the same student, caregiver, and teacher sessions that were selected for fidelity coding.

### Measures of Acceptability, Usability, Working Alliance, and Cost

Caregiver acceptability will be assessed using a 13-item caregiver satisfaction measure developed for use with HOPS (32). The *Children’s Intervention Rating Profile (CIRP),* a 7-item measure, will be adapted to assess student views of acceptability (33). Intervention provider (both school provider and research provider) usability will be examined by the *Usage Rating Profile–Intervention–Revised (URP-IR),* a 29-item measure assessing factors influencing the usage of school interventions (34). The URP-IR will only be completed once by each provider.

The working relationship between the intervention provider and student will be assessed at the end of intervention using an adaptation of the *Working Alliance Inventory-Short Version* (WAI-S; (35)). The WAI-S consists of 12 items rated on a 7-point Likert scale (1=never; 7=always) assessing agreement on tasks and goals and the student-provider bond); higher scores are more favorable. The internal consistency of this measure among middle school students was demonstrated to be.92 for intervention providers and.90 for students. In previous research, scores on the WAI-S have been shown to be associated with intervention outcomes (11). The WAI-S will be filled out by the student (“Working Alliance Inventory-Student”) and intervention provider (“Working Alliance Inventory-Counselor”).

Total costs (labor and non-labor) associated with the implementation of HOPS as well as training and technical assistance for providers in each school will be calculated according to the ingredients method (36). To do so, we will document intervention implementation activities of providers, trainers, and project assistants using time diaries, administrative records, and interviews. We will conduct time use interviews with providers at the end of each intervention period. Materials costs will be determined using acquisition costs. The costs then will be determined from the total effort on each activity and per hour wage rate of the person conducting the activity using local price levels (average salary and benefits for those in job categories matching the staff providing services). Costs for HOPS will include: (a) time to establish and maintain the relationship with each school, (b) time spent conducting the initial training for providers; (c) provider time spent implementing each student, parent, and teacher session, (d) time spent providing technical support to each provider, and (e) materials costs. We will distinguish two types of costs: cost of physical materials used for training and interventions (e.g., training manual for each provider) and costs associated with time spent for training, technical support, and intervention implementation (37). In addition, costs for initial training and tech support of providers will be differentiated from the ongoing costs of implementation to provide separate estimates of costs for start-up in each school and costs for ongoing implementation. Cost estimates will not include time and materials needed for research activities (e.g., participant recruitment and enrollment, data collection).

Intervention, training, and technical support time will be derived from administrative records of HOPS sessions, diaries completed by Dr. Langberg and technical support assistants, and training website usage data. Study staff will interview intervention providers (both school providers and research providers) after intervention to assess their experience delivering the intervention, including how much time they have spent on session preparation, delivery, and follow-up.

## Data Analyses

### Data Collection and Management

Data from caregivers, teachers, providers, and students will be collected electronically using REDCap (Research Electronic Data Capture; (38)). All participants will be assigned a unique study ID number, which will allow for a deidentified dataset at the time of analysis. Prior to conducting the main statistical analyses, the distributions of continuous variables will be checked for normality and the presence of outliers. When there are substantial deviations from normality, an appropriate method of variable transformation will be selected (such as a log transformation for skewed variables). We will use independent samples t-tests, Wilcoxon Sign Rank Test, or chi-square tests to compare groups on pre-treatment demographic characteristics (e.g., student age and gender, family socioeconomic status) and scores on outcome measures. Variables for which there are statistically significant differences between groups will be included as covariates in subsequent analyses. Continuous variables will be characterized using means, medians, ranges, and standard deviations. Categorical variables will be characterized by frequencies and percentages. Summary tables will be created that present means and standard deviations for each assessment period (Baseline, Post-Treatment, 6-Month Follow-Up, 12-Month Follow-Up) for each intervention condition, as applicable.

### Missing Data

Our team will work to keep missing data to a minimum. We will program our case report forms in REDCap to prompt respondents to answer all items if they attempt to submit the form without responding to all items. To ensure the voluntary nature of participation, we will program REDCap such that the measures can still be submitted with responses blank/missing. For all variables, we will assess the degree and pattern of missing data to see whether missing data elements can be inferred from other responses. Full information maximum likelihood (FIML) procedures will be used for any student with missing covariate information (39). FIML methods use all the observed data and produce estimates of the parameters by integrating over the missing data.

### Data and Trial Monitoring

Consistent with expectations of the study sponsor, the study did not include a data and safety monitoring committee. The confidentiality and privacy of data will be monitored carefully by the study principal investigators and statisticians on a monthly basis throughout the study. In addition, the Office of Research Compliance at the study site will conduct periodic audits to ensure compliance with federal, state, and institutional regulations. Further, the Institutional Review Board will oversee the study implementation to ensure compliance with regulations.

### Analyses for Aims 1 to 3: Evaluation of HOPS vs. TAU/WL and Comparison of HOPS-ST to HOPS-RT

*Hypothesis 1:* Students in HOPS-ST will demonstrate significantly fewer OTMP skills deficits and homework problems and will show significantly better academic performance at post-treatment compared to students in TAU-WL

*Hypothesis 2:* Students in HOPS-RT will demonstrate significantly fewer OTMP skills deficits and homework problems and will show significantly better academic performance at post-treatment compared to students in TAU-WL.

*Exploratory:* Examine whether the effects of HOPS-ST and HOPS-RT differ at post-treatment and follow-up.

A linear mixed-effects modeling approach will be used to examine intervention effects at post-treatment. An intent-to-treat approach to data analysis will be applied, including all children with baseline data in the analytic model. Under this approach, we will examine differences between students in HOPS (for HOPS-ST and HOPS-RT separately) vs. TAU-WL (as well as for students in HOPS-ST vs. HOPS-RT) by fitting a multilevel model in which students are nested within schools. In addition to including an indicator variable for the treatment condition, we will include covariates that explain variation at the student and school levels. To explore the sustainability of the intervention effect, we will examine how both means and slopes (rates of change) vary from baseline to 6-month and 12-month follow-up using regression analyses. Changes in mean and slope reflect variations in the magnitude of effect and pace of change during different phases of the study.

Because the efficacy of HOPS will be evaluated on three different domains of outcome measures (organizational skills, homework performance, academic achievement), we will use a Bonferroni-adjusted significance level to correct for multiple comparisons within each domain. As a result, the significance level will be adjusted to.025 per domain for the two measures of organizational skills (COSS-P, COSS-T), the two measures of homework performance (HPC, HPQ-T), and the two measures of academic achievement (APS, academic grades).

Power calculations were based on the following design parameters: *N= 30* schools (20 assigned to HOPS, 10 assigned to TAU); ICC = 0.04 (based on (1)); 𝛼 = 0.025 (reflecting the Bonferroni corrected significance level to accommodate multiple testing of two outcomes for each outcome domain); and 𝑅^2^ = 0.60 at the student and school level (which is the proportion of variation in outcomes explained by student and school-level covariates). Based on initial projections, we expected to enroll 8 students per school for a total of 240 participants (216 with 15% attrition), which would be sufficient to identify a minimal detectable effect size (MDES) of 0.41. Because of recruitment challenges primarily due to barriers related to the COVID-19 pandemic, we adjusted these projections to an average of 4.5 students/school for 30 schools for a total of 135 participants (122 with attrition). This sample size will result in a MDES of 0.48 with adequate power (80%). Based on prior research by Langberg and colleagues (1), the MDES for primary outcomes in our study is well within the range of expected effect sizes.

### Analyses for Aim 4: Mediation

We will use structural equation modeling (SEM) to conduct mediation analyses (40) to explre if the effect of HOPS on academic performance works through improved organizational skills. SEM approaches allow us to explore individual and multiple mediator relationships by positing a path model for each mediator variable. Applying this framework, the total intervention effect is decomposed into two components: natural direct effect (NDE = the effect of HOPS on students’ academic performance without mediation), and the natural indirect effect (NIE = the effect of HOPS that seems to proceed through improved organization skills). As a result, total effect of HOPS = NDE + NIE and a significant NIE implies that the effect of HOPS on academic performance works through improved organizational skills (41).

### Analyses for Aim 5: Effect Modification

Our plan is to explore the moderating effect of severity of organization problems, ADHD, oppositional defiant behavior, and internalizing symptoms, ADHD medication status, as well as socioeconomic level (i.e., caregiver level of education, Area Deprivation Index) at baseline on the effectiveness of HOPS. These moderators will first be examined individually in the previously described mixed effects models. Baseline scores on the moderators that are continuous variables will be centered in relation to the group-mean within each school. The moderators will be examined as a moderator by treatment group interaction term in each of the multi-level models. Significant interaction effects for both continuous and discrete variables imply moderation effects, where the magnitude and sign of the estimated coefficients reflect the moderated change in treatment effect. Following the analytic plan used by Langberg and colleagues (1), significant interaction effects among the continuous moderators will be examined at the 25^th^, 50^th^, and 75^th^ percentile levels of the moderator. For the discrete variable (medication status), we will compare treatment effects for students on and off medication at baseline to assess the magnitude of the moderating effects. In addition, we will examine the combined effect of multiple moderators. This will be done by incorporating multiple interaction terms and several higher order interaction effects in the mixed effects models (see analytic plan for Aims 1-3). The purpose of these analyses is to explore whether the inclusion and exclusion of specific moderator variables augment or attenuate the moderating effect of different variables.

Similar to Langberg and colleagues (1), we will formally test the significance of moderator effects using the models described for Aims 1-3. We will also examine the proportion of variation in the outcomes that is explained by the moderators, given by the magnitude of the adjusted R^2^ values. In addition, we will report the 95% confidence intervals for the moderator by treatment group interaction, because confidence intervals provide more precise information about the magnitude of the moderator effect (42).

### Analyses for Aim 6: Association of Fidelity, Engagement, Working Alliance to Outcomes

Our plan is to evaluate the extent to which provider intervention fidelity, student and caregiver engagement, and provider-student working alliance are associated with outcomes for participants in HOPS using multiple linear regression models. We will determine the proportion of variance in the outcome that is explained by the implementation factors, as assessed using adjusted R^2^ measures. Additionally, we will formally test the significance of the estimated coefficients to identify which implementation factor(s) are predictive of changes in the outcome measures.

### Analyses for Aim 7: Estimate of Cost

We will estimate the incremental cost of HOPS relative to TAU. Cost estimates will differentiate the cost of materials used for training and interventions and costs associated with time spent for training, technical support, and intervention implementation.

## Discussion

The main purpose of the study is to evaluate the effectiveness of the HOPS intervention when provided under authentic practice conditions by professionals working in the schools attended by enrolled students. School providers will receive training and technical support that is commonly offered when attempting to implement an evidence-based program in their schools, in contrast to more intensive consultation and coaching that is often provided through research trials (see (8)). In addition, the study will examine the effectiveness of HOPS when provided in schools by a limited number of study team members who work across multiple schools, which is similar to how the prior large-scale RCT of HOPS was conducted previously (1). The study design will enable our team to explore differences in the effectiveness of HOPS when offered by school and research team members and variations in effectiveness related to differences in intervention content and process fidelity, student-provider working alliance, and student and caregiver engagement, as well as the cost associated with intervention implementation.

Strengths of the study include the in-depth monitoring of intervention fidelity by school and research providers, including both content and process fidelity, and the assessment of student and caregiver engagement in the intervention. The assessment of school provider fidelity will provide valuable information about how HOPS is implemented in authentic practice settings, factors contributing to variable implementation, and the association of fidelity to student outcomes. Another strength of the study is the analysis of factors that may contribute to variable effects of intervention, including severity of child symptoms of ADHD, oppositional defiant behavior, internalizing symptoms, as well as family socioeconomic factors, including caregiver level of education and the level of neighborhood disadvantage for participating families (i.e., Area Deprivation Index). Additional strengths of the study are the examination of variables that might account for the impact of HOPS on child academic outcomes and the analysis of the costs of implementing the program in authentic school settings.

All students and families will be enrolled after the height of the COVID-19 pandemic when almost all schools are offering in-person instruction to students. A practical challenge is that schools will continue to experience substantial resource limitations during the period of enrollment. In the early years of the project, personnel in many schools expressed strong interest in the study but limited capacity to support research activities (e.g., implement HOPS, recruit students, contact families for initial consent, and support data collection). To be responsive to this feedback, our study team adapted by changing the research design so that only 50% of enrolled students in the HOPS condition will receive intervention from school providers (with the remaining students getting HOPS from a research provider).

A second practical challenge is that school providers often experience difficulty connecting with families to obtain their approval to be contacted by the study team to obtain caregiver consent and child assent. Our team has collaborated with school partners to identify alternative strategies for reaching out to families of referred students to obtain initial consent (e.g., draft letter for principal to send to families; flyer with QR code for families to indicate their interest in the study and share contact information).

An additional challenge is that school providers often encounter difficulties with the video recording of sessions for fidelity coding. In collaboration with school providers, we have adapted by using the video recording function available through virtual meeting platforms and providing in-person support to school providers to ensure the technology works and images of the school provider and student are being captured.

A potential limitation of the study is that each participating school is required to identify a school staff member willing and able to serve in the role of school provider. We anticipate that many schools will not have sufficient resources to assign a school professional to this role.

Therefore, the results will be generalizable to schools having the resources to identify a school professional to serve as HOPS interventionist.

We will report study findings in Clinicaltrials.gov within one year following the end of data collection. disseminate research methods and findings to researchers and practitioners in school and clinical psychology, school mental health, child and adolescent psychiatry, and pediatrics through presentations and publications in peer-reviewed journals. A research brief will be disseminated through centers of excellence at the study institution. Further, in collaboration with our institution’s Communications and Public Relations teams, we will disseminate the results and practical implications to families, teachers, intervention providers, and school administrators through press releases and notifications on our institution’s Facebook pages and X (Twitter), Blue Sky, and LinkedIn feeds. No information that could identify a participant will be included in materials disseminated to professional or lay communities.

The findings of this study will reveal the extent to which HOPS is effective under authentic practice conditions and factors that influence the impact of intervention on student outcomes. This study will also evaluate the impact of HOPS on a wide range of students with OTMP difficulties rather than just for students with ADHD. In addition, the study will identify which students are more likely to benefit from HOPS and how the intervention exerts its effect on outcomes. Importantly, the study will reveal how HOPS is implemented under real-world conditions, without intensive supervision and coaching, while identifying factors that are related to intervention implementation. Overall, the findings of this study will advance understanding of how to implement and sustain the use of evidence-based mental health and education practices in school and community settings.

### Declarations Ethics approval and consent to participate

The study was approved by the Institutional Review Board of Children’s Hospital of Philadelphia (CHOP, IRB#19-016865). Caregivers will be required to provide consent for their child to participate in the study, and students will be required to provide assent. A waiver of consent was obtained for the screening of student participants to obtain referrals and determine eligibility. Consent for the participation of teachers (who nominate students for participation and provide data on participating students) in this study will be indicated by their submitting referrals for students to participate in the study. Only authorized study staff will obtain consent from study participants. Plans to modify the study protocol must be reviewed and approved by the Institutional Review Board.

### Timelines

Data collection for this study is expected to be completed by March, 2027.

## Authors’ contributions

Each co-author has made a substantial contribution to the development and refinement of the study protocol. In addition, each co-author has had the opportunity to review the protocol and approve the final version, as described in this manuscript. JNT and TJP conceptualized the study, planned the trial design, provided oversight for the execution of the study, prepared drafts of the manuscript, and provided final approval of all manuscript edits. PFF contributed to the development of intervention fidelity and engagement measures, developed study operating procedures, provided training related to fidelity and engagement, provided oversight of research staff, and reviewed and approved the final version of the manuscript. WC and AFJ contributed to research design, provided oversight for data management and security, prepared the statistical analysis plan, contributed to the drafting of the manuscript, and reviewed and approved the final version of the manuscript. JML contributed to the development of the study protocol, provided oversight for HOPS training sessions, and reviewed and approved the final version of the manuscript. JAM developed and provided oversight for the cost assessment and analysis plan, contributed to the design of the study, and reviewed and approved the final version of the manuscript. JC, TV, BC, CD, and KLT contributed to the development and refinement of the study protocol, contributed to data collection and management, prepared or provided oversight for amendments for the IRB, and reviewed and approved the final version of this manuscript. EHW contributed to the development and refinement of the study protocol, contributed to data collection and management, contributed to the assessment of intervention fidelity, and reviewed and approved the final version of the manuscript.

## Data Availability

Deidentified research data will be made publicly available when the study is completed and published.

## Acknowledgements

We are grateful to Miranda Goodson and Aja Ingram for their contributions to the study as research assistants.

## Abbreviations

ADHD: Attention-deficit/hyperactivity disorder
APS: Academic Proficiency Scale
COSS: Children’s Organizational Skills Scale
HOPS: Homework, Organization, and Planning Skills
HPC: Homework Problem Checklist
HPQ: Homework Performance Questionnaire
ODD: Oppositional Defiant Disorder
OTMP: Organization, Time Management, and Planning
TAU/WL: Treatment-as-usual/waitlist

## Notes

### Competing Interest Statement

One of the investigators, Joshua Langberg, receives royalty payments from the National Association of School Psychologists for books about the HOPS program, which is being evaluated in this study. Thomas Power and Jennifer Mautone, investigators on this study, have a financial interest in a questionnaire (HPQ-T) being used in this study. The questionnaire was included in an appendix of a book published by Guilford Press in 2024 for which they receive royalty payments. This measure has already been evaluated and is not being evaluated or further developed in this study. Study investigators will continue to monitor competing interests and report any changes to the Office of Compliance and Privacy at CHOP.

### Clinical Trial

NCT04465708

### Funding Statement

Yes

### Author Declarations

The initial study protocol and subsequent amendments have been reviewed and approved by the IRB of Children's Hospital of Philadelphia.

